# Frailty diagnosed with the Clinical Frailty Scale stratifies the risk of covert and overt hepatic encephalopathy in patients with cirrhosis

**DOI:** 10.1101/2025.10.22.25338581

**Authors:** Shinji Unome, Takao Miwa, Sachiyo Hirata, Satomi Nakashima, Kayoko Nishimura, Mikita Oi, Masashi Aiba, Kenji Imai, Koji Takai, Masahito Shimizu

## Abstract

**Aim:** Frailty predisposes patients with cirrhosis to hepatic encephalopathy (HE). This study aimed to evaluate the effect of frailty on risk stratification for covert HE (CHE) and overt HE (OHE) in patients with cirrhosis.

**Methods:** Hospitalized patients with cirrhosis and without history of OHE were retrospectively included. Frailty was assessed using the Clinical Frailty Scale (CFS). Factors associated with CHE and OHE development were evaluated using the logistic regression and Fine–Gray competing risk regression models, respectively.

**Results:** Among 262 patients (median [interquartile range] age, 65 [55–74] years; 154 [58.8%] of female), frailty and CHE were identified in 25 (9.5%) and 82 (31.3%) patients, respectively. The prevalence of CHE was higher in patients with frailty than in those without frailty (84.0% vs. 25.7%; *p* < 0.001). During a median follow-up of 2.9 years, 40 patients (15.3%) developed OHE and 20 (7.6%) died. The incidence of OHE was higher in patients with frailty than in those without (incidence rates at 1, 3, and 5 years; 25%, 33%, and 36% vs. 5%, 11%, and 18%; *p* = 0.009). Multivariable analyses showed that CFS was an independent factor for CHE (odds ratio, 2.13; 95% confidence interval, 1.41–3.37; *p* < 0.001) and OHE development (subdistribution hazard ratio, 1.38; 95% confidence interval, 1.02–1.87; *p* = 0.037).

**Conclusions:** Frailty assessed using the CFS is a robust factor to stratify the risk of CHE and OHE development in patients with cirrhosis. Patients with frailty should be screened and carefully monitored for HE.

## Introduction

Hepatic encephalopathy (HE) is a common and devastating complication of cirrhosis, which worsens patient morbidity and survival [1]. The pathogenesis of HE is multifactorial and includes systemic inflammation, the gut-liver axis, and ammonia metabolism, leading to astrocyte swelling and cell death [1, 2]. HE demonstrates a wide spectrum of manifestations from covert HE (CHE) without specific symptoms, to overt HE (OHE), which presents as asterixis and coma. CHE is present in approximately 40% of patients with cirrhosis, and OHE develops at an annual incidence rate of 10% among patients with CHE [3, 4]. Even the mildest form of CHE is strongly associated with poor quality of life, frequent falls and accidents, and increased mortality [4, 5]. When CHE progresses to OHE, patients and their caregivers experience impaired social functioning, frequent hospitalizations, and economic burden related to treatment [5]. Therefore, the early detection of CHE and accurate prediction of OHE are essential for optimizing clinical management and reducing the burden on patients with cirrhosis. Neuropsychological tests with test batteries are the reference standard for identifying CHE; however, the time requirement, need for trained practitioners, and associated costs limit their clinical use. Furthermore, even simplified tools such as the Stroop test and animal naming test are not performed in all patients with cirrhosis, reflecting limited medical resources available in real-world clinical practice [6]. Therefore, developing an accessible and simple method for stratifying the risk of CHE and OHE is essential for establishing effective screening for CHE and careful monitoring of OHE development.

Frailty is a multidimensional syndrome characterized by reduced physiological reserves across the physical, functional, and cognitive domains [7, 8]. In patients with cirrhosis, the prevalence of frailty varies by assessment method, ranging from 17% to 43% [9, 10]. Patients with frailty, particularly those awaiting liver transplantation, have an increased risk of adverse outcomes, including liver decompensation, hospitalization, and mortality [9-11]. The concept of frailty emphasizes on muscle dysfunction, reduced mobility, functional dependence, and cognitive vulnerability, all of which are strongly linked to CHE and OHE [8]. Therefore, the diagnosis of frailty can capture high-risk populations and serve as an effective method for identifying those at risk for both CHE and OHE. The Clinical Frailty Scale (CFS) is a brief, global frailty assessment tool initially developed for use in the geriatric population [12, 13]. The CFS has also been validated in patients with cirrhosis and is, therefore, recommended by the American Association for the Study of Liver Diseases as a reliable method for identifying frailty [8, 14, 15]. The strengths of the CFS include minimal training requirements, short assessment time, and no necessity for specialized equipment, all of which allow it to be used by non-specialists in both inpatient and outpatient settings.

Given this background, we hypothesized that frailty assessed using the CFS could help identify patients with cirrhosis at a higher risk of developing HE. This study aimed to evaluate the utility of the CFS in stratifying the risk of CHE and OHE development in patients with cirrhosis.

## Materials and Methods

### Study design

This retrospective observational study enrolled patients with cirrhosis who were hospitalized at Gifu University Hospital. The study protocol was reviewed and approved by the Ethics Committee of Gifu University School of Medicine (approval number: 2024-270). This study adhered to the ethical guidelines of the Declaration of Helsinki. Given the retrospective nature of the study, informed consent was obtained using an opt-out approach.

### Participants

This study included hospitalized patients with cirrhosis of any etiology, without a history of OHE or hepatocellular carcinoma, aged ≥18 years, and who underwent CHE assessment at Gifu University Hospital between March 3rd 2004 and December 6th 2023. Cirrhosis was diagnosed based on the clinical presentation, laboratory data, imaging findings, and liver histology. The exclusion criteria were as follows: history of organ transplantation; active malignancy; neurological or psychiatric disorders; acute systemic diseases, including acute hepatitis, acute-on-chronic liver failure, bacterial infection, ruptured varices; life-threatening comorbidities; large portosystemic shunts; transjugular intrahepatic portosystemic shunt; and opt-out refusal. All patients received standard medical care in accordance with the guidelines for cirrhosis [16, 17] and were followed up until the last visit, death, or August 21, 2024, whichever occurred first.

### Outcomes and data collection

The study endpoint was the association between frailty and HE, including the presence of CHE and development of OHE in patients with cirrhosis.

Baseline data were collected from medical records within 1 week of admission and included the following variables: age, sex, body mass index, etiology of cirrhosis, presence of ascites and CHE, and laboratory findings. The etiology of cirrhosis was categorized as viral hepatitis, alcohol-related liver disease, metabolic dysfunction-associated steatotic liver disease, or other causes. Ascites was identified using medical imaging. The body mass index was calculated using the estimated dry weight, adjusted by subtracting the percentage of measured weight based on fluid retention: 5% for mild ascites, 10% for moderate ascites, 15% for severe ascites, and an additional 5% for bilateral pedal edema [18]. The authors did not have access to any information that could identify individual participants during or after data collection. All data were anonymized before transfer to the principal investigator.

### Diagnosis of CHE and OHE

CHE was diagnosed using a computer-aided neuropsychiatric test (NPT) conducted within 1 week of admission by trained practitioners, using standardized software (Otsuka Pharmaceutical Co., Ltd., Tokyo, Japan). The NPT comprises four subtests: the number connection tests A and B, digit symbol test, and block design test. CHE was diagnosed when patients exhibited abnormalities in two or more of the four subtests [19, 20]. The NPT is recommended by the Japan Society of Hepatology as the gold-standard tool for diagnosing CHE and has been validated as a predictor of OHE in Japanese patients with cirrhosis [20, 21]. OHE diagnosis was based on clinical assessment using the West Haven criteria [5].

### Diagnosis of frailty

Frailty was retrospectively assessed using the CFS, based on a structured questionnaire of admission-day medical records, including comorbidities, activities of daily living, and fall risk. The CFS comprises nine categories based on the patient’s condition (1 = very fit; 2 = fit; 3 = managing well; 4 = very mild frailty; 5 = mild frailty; 6 = moderate frailty; 7 = severe frailty; 8 = very severe frailty; and 9 = terminally ill) [12, 13]. Frailty was defined as a CFS > 4 (CFS 5–9), in line with previously validated criteria [13].

### Statistical analyses

Baseline characteristics were summarized as numbers with percentages for categorical variables and as medians with interquartile ranges for continuous variables. Comparisons between groups were performed using the chi-square test for categorical variables and the Mann–Whitney *U* test for continuous variables. Factors associated with the presence of CHE were analyzed using a multivariable logistic regression model, with the results reported as odds ratios (ORs) and 95% confidence intervals (CIs). Considering death as a competing event, the cumulative incidence curves of OHE were estimated using the cumulative incidence function and compared using the Gray’s test. In addition, factors associated with OHE occurrence were investigated using the Fine–Gray competing risk regression model, with the results reported as subdistribution hazard ratios (SHRs) with 95% CIs. All statistical tests were two-sided, with *p-*values < 0.05 considered statistically significant. Statistical analyses were performed using the R software version 4.4.0 (R Foundation for Statistical Computing, Vienna, Austria).

## Results

### Baseline characteristics of patients with cirrhosis

Among the 393 screened patients, 262 met the eligibility criteria and were included in the analysis (S1 Fig). The baseline clinical characteristics of the patients are shown in Table 1. The median age was 65 years, 154 (58.8%) patients were female, and the median body mass index was 22.3 kg/m^2^. The most common etiologies of cirrhosis were alcohol-related liver disease (24.8%), viral hepatitis (22.5%), and metabolic dysfunction-associated steatotic liver disease (18.7%). The median Child–Pugh, model for end-stage liver disease (MELD) and albumin-bilirubin (ALBI) scores were 6, 8 and −2.28, respectively. The median CFS score for the entire cohort was 3. Based on the NPT, CHE was observed in 82 (31.3%) patients.

**Table 1.** Baseline characteristics of patients with cirrhosis divided by the presence of frailty.

| Characteristic | Overall<br>(n = 262) | No frailty<br>(n = 237) | Frailty<br>(n = 25) | *p*-value* |
| --- | --- | --- | --- | --- |
| Age (years) | 65 [55–74] | 64 [55–73] | 71 [61–75] | <0.001 |
| Female sex | 154 (58.8) | 140 (59.1) | 14 (56.0) | 0.934 |
| Body mass index (kg/m<sup>2</sup>) | 22.3 [19.8–25.6] | 22.6 [20.1–25.8] | 19.6 [18.7–22.5] | 0.001 |
| Etiology |  |  |  | 0.425 |
| Viral | 59 (22.5) | 53 (22.4) | 6 (24.0) |  |
| ALD | 65 (24.8) | 57 (24.1) | 8 (32.0) |  |
| MASLD | 49 (18.7) | 45 (19.0) | 4 (16.0) |  |
| Others | 89 (34.0) | 82 (34.6) | 7 (28.0) |  |
| Ascites | 99 (37.8) | 78 (34.0) | 21 (84.0) | <0.001 |
| Varices | 110 (42.2) | 96 (40.6) | 14 (56.0) | 0.238 |
| Child–Pugh class |  |  |  | <0.001 |
| A | 145 (55.3) | 144 (60.8) | 1 (4.0) |  |
| B | 68 (26.0) | 60 (25.3) | 8 (32.0) |  |
| C | 49 (18.7) | 33 (13.9) | 16 (64.0) |  |
| Child–Pugh score | 6 [5–9] | 6 [5–8] | 11 [8–11] | <0.001 |
| MELD score | 8 [7–11] | 8 [7–10] | 11 [9–16] | <0.001 |
| ALBI score | -2.28 [-2.67, -1.50] | -2.35 [-2.70, -1.58] | -1.03 [-1.58, -0.57] | <0.001 |
| Albumin (g/dL) | 3.6 [2.9–4.0] | 3.7 [3.0–4.1] | 2.3 [2.1–2.7] | <0.001 |
| Bilirubin (mg/dL) | 1.1 [0.8–1.6] | 1.1 [0.8–1.6] | 1.6 [0.9–2.6] | 0.026 |
| Creatinine (mg/dL) | 0.67 [0.54–0.80] | 0.66 [0.54–0.80] | 0.79 [0.64–1.05] | 0.004 |
| Sodium (mEq/L) | 139 [137–140] | 139 [137–140] | 136 [133–138] | <0.001 |
| Platelet (10<sup>9</sup>/L) | 119 [86–188] | 124 [86–188] | 102 [73–158] | 0.363 |
| International normalized ratio | 1.06 [0.99–1.22] | 1.05 [0.98–1.17] | 1.23 [1.07–1.42] | <0.001 |
| Ammonia (μg/dL) | 56 [39–79] | 57 [39–79] | 49 [37–80] | 0.772 |
| CHE | 82 (31.3) | 61 (25.7) | 21 (84.0) | <0.001 |
| Clinical Frailty Scale | 3 [3–3] | 3 [3–3] | 6 [5–6] | <0.001 |

Values are presented as number (percentage) or median (interquartile range).
\*Groups were compared using the chi-square test or Mann–Whitney *U* test.
Abbreviations: ALBI, albumin-bilirubin; ALD, alcohol-associated/related disease; CHE, covert hepatic encephalopathy; MASLD, metabolic dysfunction-associated steatotic liver disease; MELD, model for end-stage liver disease

### Comparisons between patients with and without frailty

Frailty was identified in 25 (9.5%) patients with a median CFS score of 6 points. Comparisons between patients with and without frailty are presented in Table 1. Compared with patients without frailty, those with frailty had more advanced liver disease, as indicated by higher Child–Pugh, MELD, and ALBI scores, and differences in ascites, albumin, bilirubin, creatinine, sodium, and international normalized ratio. Additionally, patients with frailty had a significantly higher prevalence of CHE than those without frailty (84.0 vs. 25.7%; *p* < 0.001; Fig 1).

**Fig 1.**
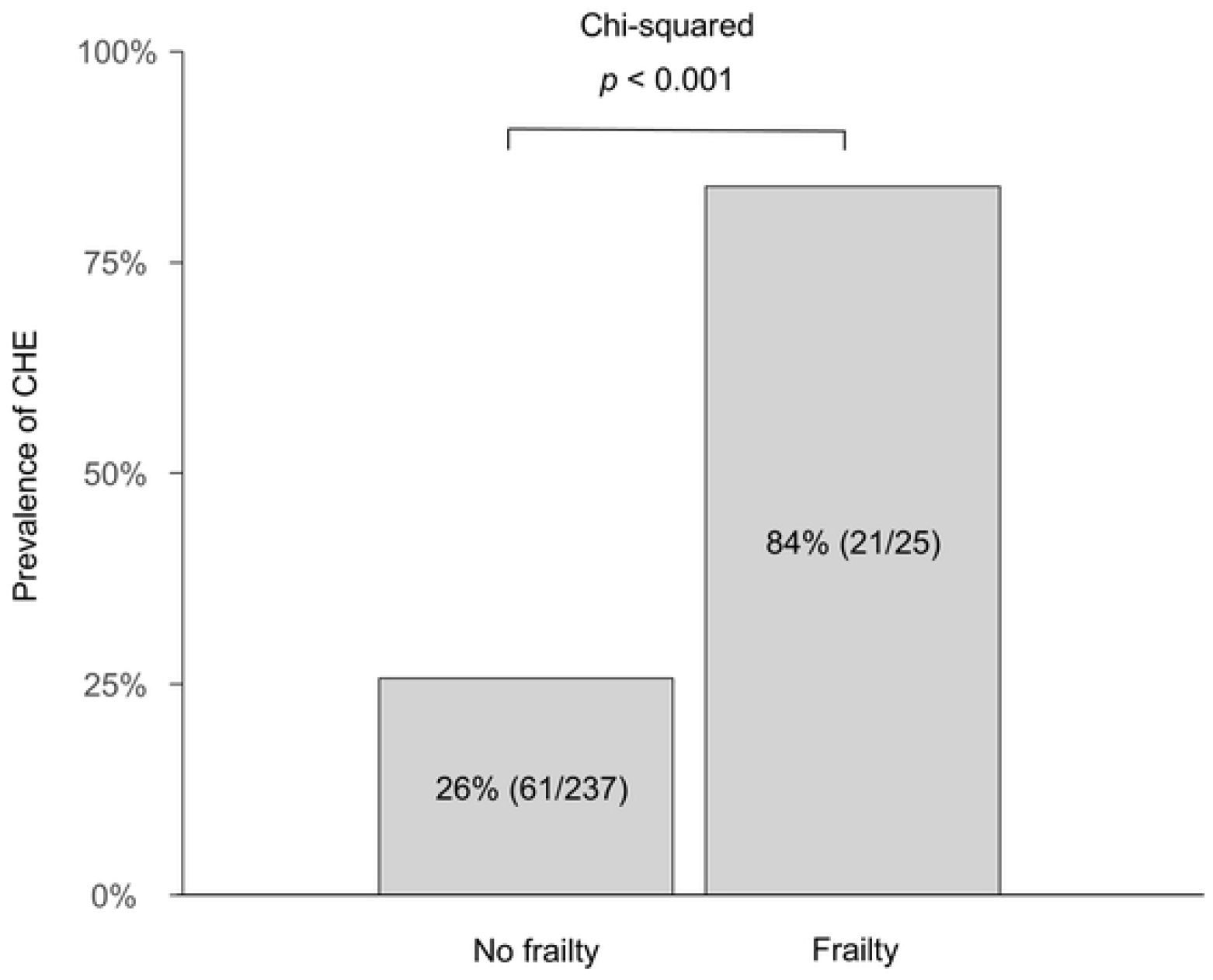
Prevalence of covert hepatic encephalopathy in patients with cirrhosis according to the presence of frailty.

### Impact of frailty on the presence of CHE in patients with cirrhosis

Independent factors associated with the presence of CHE in patients with cirrhosis are shown in Table 2. After adjustment, multivariable model 1 identified that CFS (OR, 2.13; 95% CI, 1.41–3.37; *p* < 0.001) and Child–Pugh score (OR, 1.37; 95% CI, 1.19–1.58; *p* < 0.001) were independently associated with CHE. Similar associations were observed in models 2 and 3, which included the MELD and ALBI scores instead of the Child–Pugh score, respectively.

**Table 2.**
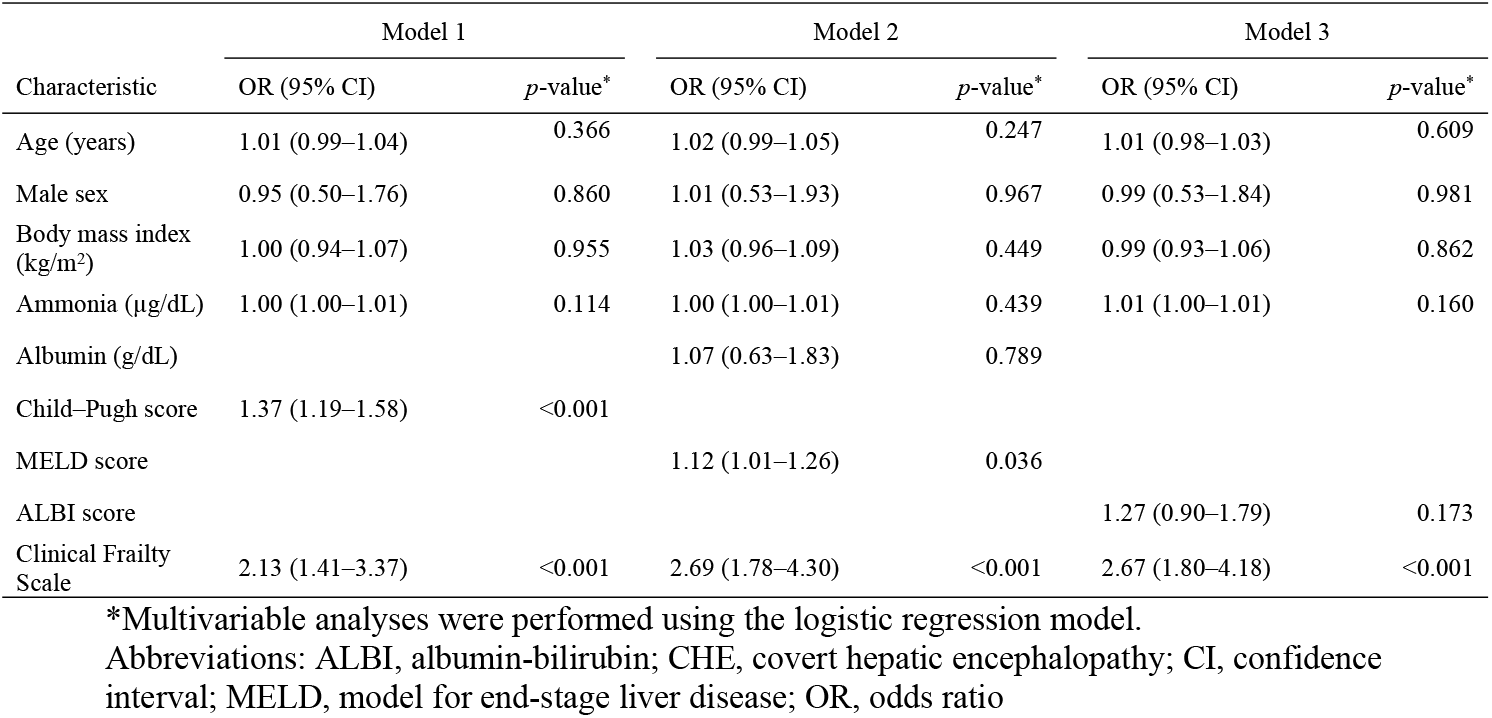
Impact of frailty on CHE in patients with cirrhosis.

| Characteristic | Model 1 |  | Model 2 |  | Model 3 |  |
| --- | --- | --- | --- | --- | --- | --- |
|  | OR (95% CI) | *p*-value* | OR (95% CI) | *p*-value* | OR (95% CI) | *p*-value* |
| Age (years) | 1.01 (0.99–1.04) | 0.366 | 1.02 (0.99–1.05) | 0.247 | 1.01 (0.98–1.03) | 0.609 |
| Male sex | 0.95 (0.50–1.76) | 0.860 | 1.01 (0.53–1.93) | 0.967 | 0.99 (0.53–1.84) | 0.981 |
| Body mass index (kg/m<sup>2</sup>) | 1.00 (0.94–1.07) | 0.955 | 1.03 (0.96–1.09) | 0.449 | 0.99 (0.93–1.06) | 0.862 |
| Ammonia (μg/dL) | 1.00 (1.00–1.01) | 0.114 | 1.00 (1.00–1.01) | 0.439 | 1.01 (1.00–1.01) | 0.160 |
| Albumin (g/dL) |  |  | 1.07 (0.63–1.83) | 0.789 |  |  |
| Child–Pugh score | 1.37 (1.19–1.58) | <0.001 |  |  |  |  |
| MELD score |  |  | 1.12 (1.01–1.26) | 0.036 |  |  |
| ALBI score |  |  |  |  | 1.27 (0.90–1.79) | 0.173 |
| Clinical Frailty Scale | 2.13 (1.41–3.37) | <0.001 | 2.69 (1.78–4.30) | <0.001 | 2.67 (1.80–4.18) | <0.001 |

\*Multivariable analyses were performed using the logistic regression model.
Abbreviations: ALBI, albumin-bilirubin; CHE, covert hepatic encephalopathy; CI, confidence interval; MELD, model for end-stage liver disease; OR, odds ratio

### Impact of frailty on the development of OHE in patients with cirrhosis

During the median follow-up of 2.8 years (interquartile range, 0.8–5.5 years), 40 patients (15.3%) developed OHE, and 20 (7.6%) died. The cumulative incidences of OHE at 1, 3, and 5 years were 7%, 13%, and 20%, respectively in the overall cohort. Patients with frailty had a significantly higher incidence of OHE than those without frailty (median incidence rates at 1, 3, and 5 years; 25%, 33%, and 36% vs. 5%, 11%, and 18%, respectively; *p* = 0.009; Fig 2). Multivariable competing risk regression model 1 showed that the CFS score was an independent predictor of OHE development (SHR 1.38; 95% CI 1.02–1.87; *p* = 0.037; Table 3). Similar findings were observed in models 2 and 3.

**Table 3.** Impact of frailty on OHE development in patients with cirrhosis.

| Characteristic | Model 1 |  | Model 2 |  | Model 3 |  |
| --- | --- | --- | --- | --- | --- | --- |
|  | SHR (95% CI) | *p*-value* | SHR (95% CI) | *p*-value* | SHR (95% CI) | *p*-value* |
| Age (years) | 1.03 (1.00–1.07) | 0.069 | 1.04 (1.00–1.07) | 0.056 | 1.02 (0.99–1.05) | 0.300 |
| Male sex | 0.81 (0.39–1.67) | 0.565 | 0.66 (0.32–1.37) | 0.267 | 0.78 (0.37–1.67) | 0.522 |
| Body mass index (kg/m<sup>2</sup>) | 1.00 (0.95–1.06) | 0.928 | 1.00 (0.94–1.07) | 0.965 | 1.00 (0.94–1.07) | 0.972 |
| Ammonia (μg/dL) | 1.01 (1.01–1.02) | <0.001 | 1.01 (1.01–1.02) | <0.001 | 1.01 (1.00–1.02) | 0.008 |
| Albumin (g/dL) |  |  | 0.70 (0.39–1.28) | 0.246 |  |  |
| Child–Pugh score | 1.52 (1.33–1.75) | <0.001 |  |  |  |  |
| MELD score |  |  | 1.20 (1.08–1.33) | <0.001 |  |  |
| ALBI score |  |  |  |  | 2.18 (1.57–3.02) | <0.001 |
| CHE | 0.80 (0.36–1.78) | 0.584 | 1.07 (0.49–2.31) | 0.868 | 1.41 (0.67–3.00) | 0.367 |
| Clinical Frailty Scale | 1.38 (1.02–1.87) | 0.037 | 1.51 (1.12–2.03) | 0.006 | 1.39 (1.00–1.92) | 0.047 |

\*Multivariable analyses were performed using the Fine-Gray model.
Abbreviations: ALBI, albumin-bilirubin; CHE, covert hepatic encephalopathy; CI, confidence interval; SHR, subdistribution hazard ratio; MELD, model for end-stage liver disease; OHE, overt hepatic encephalopathy

**Fig 2.**
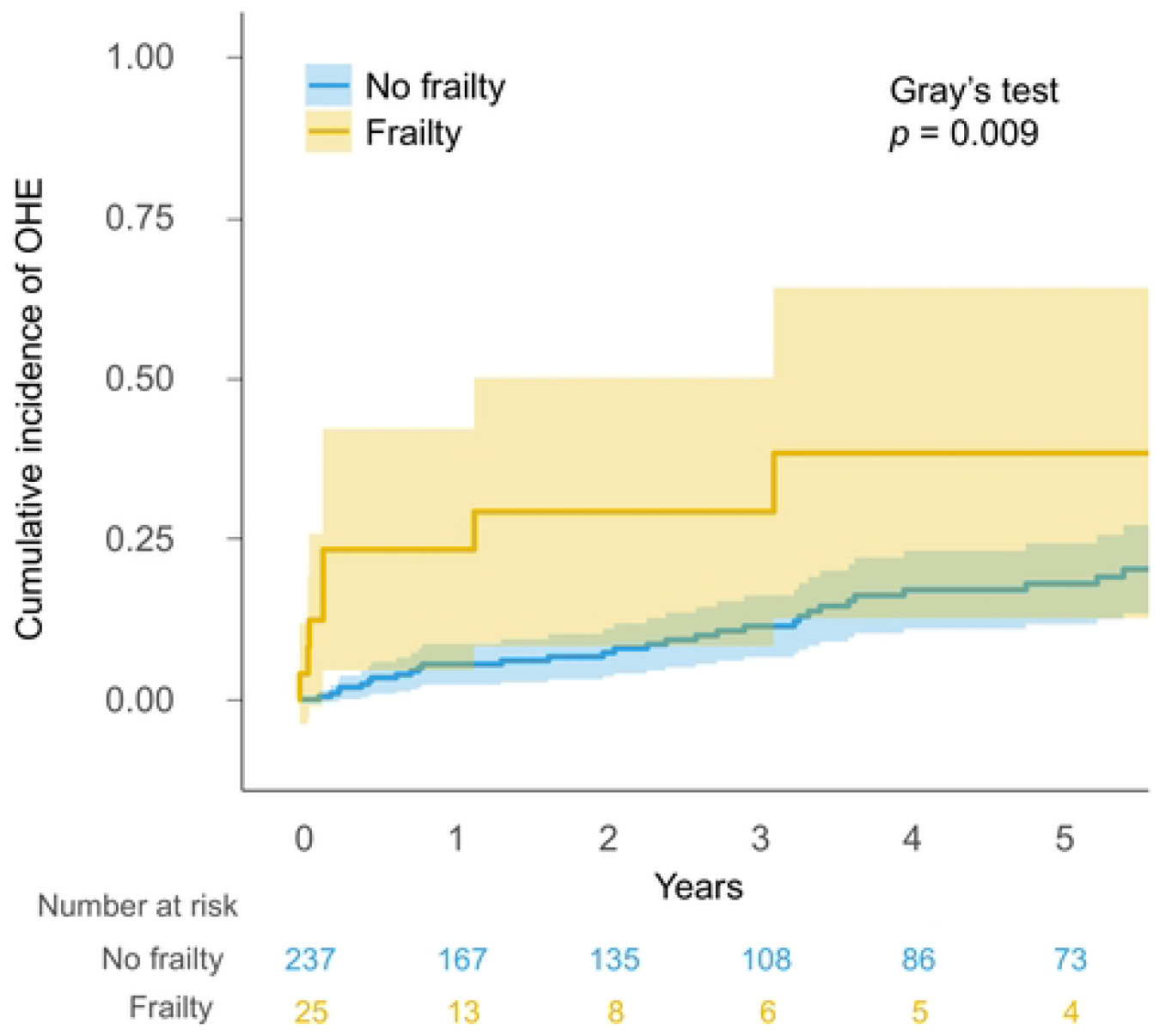
Cumulative incidence of overt hepatic encephalopathy development in patients with cirrhosis according to the presence of frailty.

## Discussion

The early detection of CHE and accurate prediction of OHE are crucial for the management of cirrhosis. Therefore, simple and easy-to-implement tools are urgently required in daily practice. In this study, frailty, assessed using the CFS, detected the presence of CHE. In addition, frailty significantly stratified the risk of developing OHE. Notably, this is the first real-world study to demonstrate the utility of the CFS in predicting both CHE and OHE in Japanese patients with cirrhosis.

The first key finding is the robust association between frailty and CHE. Physical frailty involves muscle depletion and decreased muscle strength, impairing the ammonia detoxification capacity. The influence of these effects on CHE were previously highlighted in several studies [22, 23]. Beyond muscle loss, frailty reflects a broader construct encompassing not only physical decline but also cognitive and functional vulnerability, which may predispose patients to HE [24]. These factors may explain why frailty, which integrates multiple domains of vulnerability, was a significant predictor of CHE in our cohort. To date, only a few studies have explored the relationship between CFS and CHE. A study in Germany reported that pre-frailty, as defined by the CFS, was associated with an increased likelihood of CHE diagnosed using the portosystemic encephalopathy syndrome test and animal naming test, in patients with and without prior OHE [25]. Studies using physical performance-based metrics, such as handgrip strength and composite frailty scores, have also linked frailty to CHE [23, 26, 27]. Our findings are consistent with these previous reports and further strengthen the evidence by assessing frailty with the CFS, a validated method for diagnosing frailty; diagnosing CHE using the NPT, a guideline-recommended diagnostic modality for CHE; and employing the largest cohort of CHE assessments in Japan. Notably, a recent Japanese nationwide survey on HE reported that over 90% of physicians in caring for patients with cirrhosis had not tested even half of their patients, highlighting the critical need to identify the population for whom testing is truly necessary owing to the high risk of CHE [6]. In this context, owing to its simplicity, the CFS is suitable for routine implementation, including in resource-limited or community settings, and enables the identification of patients at risk of CHE who might otherwise remain unrecognized.

Another key finding was that frailty assessed using the CFS was a robust predictor of OHE development in patients with cirrhosis. Notably, the CFS remained statistically significant even after adjusting for CHE. The independence of CFS and CHE in the development of OHE suggests that frailty may influence the risk of OHE through mechanisms other than those associated with CHE. Patients with frailty may be more vulnerable to common risk factors for OHE development, such as infection, dehydration, and constipation, due to impaired physiological reserve, nutritional deficits, and reduced self-care capacity [28-30]. In fact, previous studies have demonstrated that patients with frailty have a higher risk of a composite outcome of liver decompensation than those without frailty [31]. In addition, a prospective cohort study involving 355 patients with cirrhosis demonstrated that a combined score incorporating the CFS and Montreal Cognitive Assessment could stratify the risk of HE-related hospitalization within 6 months [32]. Furthermore, measurement of handgrip strength, an objective assessment tool for frailty, has also been identified as a predictor of OHE [23]. Our findings, together with the aforementioned reports, reveal the robustness of the association between frailty measurements and OHE development. This is also supported by a German study which found that the CFS predicted OHE occurrence regardless of baseline CHE status [25]. Our findings expand the existing knowledge between OHE development and frailty assessed by the CFS, using a large Japanese cohort and robust analysis with adjustment for confounders. Therefore, incorporating frailty assessment into routine clinical evaluation may allow earlier identification of patients at high-risk of OHE who require careful observations and timely intervention.

Certain limitations must be considered in this study. First, the retrospective design may have introduced selection bias and unmeasured confounders. Second, because this was a single-center study of hospitalized Japanese patients, the results may not be generalizable to other regions or populations. However, this study also has several strengths, including the use of a validated global frailty measure, guideline-recommended ascertainment of CHE, evaluation of both CHE and OHE using competing-risk methods in a relatively large real-world cohort, and multivariable adjustment for major confounders. Future prospective studies should examine whether interventions targeting frailty can reduce the risk of CHE and OHE and improve long-term outcomes in patients with cirrhosis.

In conclusion, frailty, as assessed by the CFS, was a robust predicter of both CHE presence and OHE development. This simple and practical assessment tool may support the early detection of CHE and identification of patients with cirrhosis at high-risk of OHE.

## Data Availability

The data and codes used in this study are available from the corresponding author upon request. However, additional approval from the Ethics Review Committee of the Graduate School of Medicine, Gifu University is required to share the data, in accordance with Japanese ethical guidelines.

## Acknowledgments

We would like to thank all the medical staff involved in healthcare at Gifu University Hospital.

## Supporting Information

**S1 Fig**. Flow diagram of the study

## Notes

### Competing Interest Statement

The authors have declared no competing interest.

### Funding Statement

Yes

### Author Declarations

The study protocol was reviewed and approved by the Ethics Committee of Gifu University School of Medicine (approval number: 2024-270). This study adhered to the ethical guidelines of the Declaration of Helsinki.

## References

1. Fallahzadeh MA, Rahimi RS. Hepatic Encephalopathy: Current and Emerging Treatment Modalities. Clin Gastroenterol Hepatol. 2022;20:S9–S19. doi:10.1016/j.cgh.2022.04.034

2. Rose CF, Amodio P, Bajaj JS, et al. Hepatic encephalopathy: Novel insights into classification, pathophysiology and therapy. J Hepatol. 2020;73:1526–47. doi:10.1016/j.jhep.2020.07.013

3. Lv XH, Lu Q, Deng K, et al. Prevalence and Characteristics of Covert/Minimal Hepatic Encephalopathy in Patients With Liver Cirrhosis: A Systematic Review and Meta-Analysis. Am J Gastroenterol. 2024;119:690–9. doi:10.14309/ajg.0000000000002563

4. European AftSotL. EASL Clinical Practice Guidelines on the management of hepatic encephalopathy. J Hepatol. 2022;77:807–24. doi:10.1016/j.jhep.2022.06.001

5. Vilstrup H, Amodio P, Bajaj J, et al. Hepatic encephalopathy in chronic liver disease: 2014 Practice Guideline by the American Association for the Study of Liver Diseases and the European Association for the Study of the Liver. Hepatology. 2014;60:715–35. doi:10.1002/hep.27210

6. Miwa T, Tsuruoka M, Ueda H, et al. Current management and future perspectives of covert hepatic encephalopathy in Japan: a nationwide survey. J Gastroenterol. 2025. doi:10.1007/s00535-025-02232-0

7. Kim DH, Rockwood K. Frailty in Older Adults. N Engl J Med. 2024;391:538–48. doi:10.1056/NEJMra2301292

8. Lai JC, Tandon P, Bernal W, et al. Malnutrition, Frailty, and Sarcopenia in Patients With Cirrhosis: 2021 Practice Guidance by the American Association for the Study of Liver Diseases. Hepatology. 2021;74:1611–44. doi:10.1002/hep.32049

9. Padhi BK, Gandhi AP, Sandeep M, et al. Prevalence of Frailty and Its Impact on Mortality and Hospitalization in Patients With Cirrhosis: A Systematic Review and Meta-analysis. J Clin Exp Hepatol. 2024;14:101373. doi:10.1016/j.jceh.2024.101373

10. Laube R, Wang H, Park L, et al. Frailty in advanced liver disease. Liver Int. 2018;38:2117–28. doi:10.1111/liv.13917

11. Lai JC, Rahimi RS, Verna EC, et al. Frailty Associated With Waitlist Mortality Independent of Ascites and Hepatic Encephalopathy in a Multicenter Study. Gastroenterology. 2019;156:1675–82. doi:10.1053/j.gastro.2019.01.028

12. Rockwood K, Theou O. Using the Clinical Frailty Scale in Allocating Scarce Health Care Resources. Can Geriatr J. 2020;23:210–5. doi:10.5770/cgj.23.463

13. Rockwood K, Song X, MacKnight C, et al. A global clinical measure of fitness and frailty in elderly people. CMAJ. 2005;173:489–95. doi:10.1503/cmaj.050051

14. Kremer WM, Nagel M, Reuter M, et al. Validation of the Clinical Frailty Scale for the Prediction of Mortality in Patients With Liver Cirrhosis. Clin Transl Gastroenterol. 2020;11:e00211. doi:10.14309/ctg.0000000000000211

15. Tandon P, Tangri N, Thomas L, et al. A Rapid Bedside Screen to Predict Unplanned Hospitalization and Death in Outpatients With Cirrhosis: A Prospective Evaluation of the Clinical Frailty Scale. Am J Gastroenterol. 2016;111:1759–67. doi:10.1038/ajg.2016.303

16. Yoshiji H, Nagoshi S, Akahane T, et al. Evidence-based clinical practice guidelines for liver cirrhosis 2020. Hepatol Res. 2021;51:725–49. doi:10.1111/hepr.13678

17. Yoshiji H, Nagoshi S, Akahane T, et al. Evidence-based clinical practice guidelines for Liver Cirrhosis 2020. J Gastroenterol. 2021;56:593–619. doi:10.1007/s00535-021-01788-x

18. European AftSotL. EASL Clinical Practice Guidelines on nutrition in chronic liver disease. J Hepatol. 2019;70:172–93. doi:10.1016/j.jhep.2018.06.024

19. Kato A, Watanabe Y, Sawara K, et al. Diagnosis of sub-clinical hepatic encephalopathy by Neuropsychological Tests (NP-tests). Hepatol Res. 2008;38 Suppl 1:S122–7. doi:10.1111/j.1872-034X.2008.00437.x

20. Kawaguchi T, Konishi M, Kato A, et al. Updating the neuropsychological test system in Japan for the elderly and in a modern touch screen tablet society by resetting the cut-off values. Hepatol Res. 2017;47:1335–9. doi:10.1111/hepr.12864

21. Miwa T, Hanai T, Nishimura K, et al. A simple covert hepatic encephalopathy screening model based on blood biochemical parameters in patients with cirrhosis. PLoS One. 2022;17:e0277829. doi:10.1371/journal.pone.0277829

22. Merli M, Giusto M, Lucidi C, et al. Muscle depletion increases the risk of overt and minimal hepatic encephalopathy: results of a prospective study. Metab Brain Dis. 2013;28:281–4. doi:10.1007/s11011-012-9365-z

23. Miwa T, Hanai T, Nishimura K, et al. Handgrip strength stratifies the risk of covert and overt hepatic encephalopathy in patients with cirrhosis. JPEN J Parenter Enteral Nutr. 2022;46:858–66. doi:10.1002/jpen.2222

24. Berry K, Duarte-Rojo A, Grab JD, et al. Cognitive Impairment and Physical Frailty in Patients With Cirrhosis. Hepatol Commun. 2022;6:237–46. doi:10.1002/hep4.1796

25. Schleicher EM, Kaps L, Schattenberg JM, et al. Higher scores in the Clinical Frailty Scale are associated with covert and overt hepatic encephalopathy in patients with cirrhosis. Dig Liver Dis. 2024;56:1046–53. doi:10.1016/j.dld.2023.12.001

26. San Martin-Valenzuela C, Borras-Barrachina A, Gallego JJ, et al. Motor and Cognitive Performance in Patients with Liver Cirrhosis with Minimal Hepatic Encephalopathy. J Clin Med. 2020;9. doi:10.3390/jcm9072154

27. Sogbe M, Wong R, Bloomer PM, et al. Psychomotor Speed From Minimal Hepatic Encephalopathy Testing Is Associated With Physical Frailty in Patients With Advanced Chronic Liver Disease. Am J Gastroenterol. 2025. doi:10.14309/ajg.0000000000003583

28. Yang Y, Che K, Deng J, et al. Assessing the Impact of Frailty on Infection Risk in Older Adults: Prospective Observational Cohort Study. JMIR Public Health Surveill. 2024;10:e59762. doi:10.2196/59762

29. Lim J, Park H, Lee H, et al. Higher frailty burden in older adults with chronic constipation. BMC Gastroenterol. 2021;21:137. doi:10.1186/s12876-021-01684-x

30. Schleicher EM, Kremer WM, Kalampoka V, et al. Frailty as Tested by the Clinical Frailty Scale Is a Risk Factor for Hepatorenal Syndrome in Patients With Liver Cirrhosis. Clin Transl Gastroenterol. 2022;13:e00512. doi:10.14309/ctg.0000000000000512

31. Alvares-da-Silva MR, Reverbel da Silveira T. Comparison between handgrip strength, subjective global assessment, and prognostic nutritional index in assessing malnutrition and predicting clinical outcome in cirrhotic outpatients. Nutrition. 2005;21:113–7. doi:10.1016/j.nut.2004.02.002

32. Ney M, Tangri N, Dobbs B, et al. Predicting Hepatic Encephalopathy-Related Hospitalizations Using a Composite Assessment of Cognitive Impairment and Frailty in 355 Patients With Cirrhosis. Am J Gastroenterol. 2018;113:1506–15. doi:10.1038/s41395-018-0243-0

